# Estimating the relative proportions of SARS-CoV-2 strains from wastewater samples

**DOI:** 10.1101/2022.01.13.22269236

**Authors:** Lenore Pipes, Zihao Chen, Svetlana Afanaseva, Rasmus Nielsen

## Abstract

Wastewater surveillance has become essential for monitoring the spread of SARS-CoV-2. The quantification of SARS-CoV-2 RNA in wastewater correlates with the Covid-19 caseload in a community. However, estimating the proportions of different SARS-CoV-2 strains has remained technically difficult. We present a method for estimating the relative proportions of SARS-CoV-2 strains from wastewater samples. The method uses an initial step to remove unlikely strains, imputation of missing nucleotides using the global SARS-CoV-2 phylogeny, and an Expectation-Maximization (EM) algorithm for obtaining maximum likelihood estimates of the proportions of different strains in a sample. Using simulations with a reference database of >3 million SARS-CoV-2 genomes, we show that the estimated proportions accurately reflect the true proportions given sufficiently high sequencing depth and that the phylogenetic imputation is highly accurate and substantially improves the reference database.

## Introduction

The ongoing pandemic of coronavirus disease of 2019 (Covid-19) caused by severe acute respiratory syndrome coronavirus 2 (SARS-CoV-2) continues to be the world’s worst public health emergency in the last century. There is an emerging need to identify the initiation of outbreaks, distribution, and changing trends of Covid-19 in near real-time (***Korber et al., 2020***; ***Rockett et al., 2020***). Wastewater-based epidemiology (WBE) has become an effective monitoring strategy for early detection of SARS-CoV-2 in communities as well as being an important method for informing public health interventions aimed at containing and mitigating Covid-19 outbreaks (***Ahmed et al., 2020***). WBE for SARS-CoV-2 can detect the virus excreted by both symptomatic and asymptomatic individuals alike thus making it an effective approach for modeling the disease signature of entire communities. WBE data also strongly correlates with the Covid-19 case rates in the community (***Medema et al., 2020a***; ***Farkas et al., 2020***).

Currently, most analyses of WBE data for SARS-CoV-2 focus on identifying presence/absence as well as quantifying the abundance of the virus (***Kumar et al., 2020***; ***Crits-Christoph et al., 2021***; ***Wu et al., 2020***; ***Medema et al., 2020b***). However, identifying and profiling multiple SARS-CoV-2 genotypes in a single sample can provide additional information for understanding the dynamics and transmission of certain strains. The alarming continued emergence of novel variants such as the Delta variant, B.1.617.2, and the Omicron variant, B.1.1.529, underscores the urgency and need for quantification of the abundance of different viral strains across communities. Unfortunately, it is difficult to precisely quantify the proportions of different strains of a virus in an environmental sample, such as wastewater, using standard sequencing technologies given the low quality and highly uneven depth of sequencing data. Adding to these challenges is that many strains are nearly identical differing by only one or a few mutations across approximately ∼30,000 nucleotides. With millions of possible candidate strain the combinatorial challenge of identifying the correct strains is large, particularly when strains are not identified by individual diagnostic mutations, but rather by sets of mutations that jointly helps distinguish the strains from each other. Nonetheless, quantification of strain composition in WBE data has the potential to become a cost-effective method to identify changes in viral community composition as SARS-CoV-2 becomes an endemic virus.

We present a method for estimating the proportion of different SARS-CoV-2 strains from shot-gun sequencing of wastewater samples allowing researchers to obtain results in real-time. The method is based on an initial filtering step, phylogenetic imputation of missing nucleotides, and an Expectation-Maximization (EM) algorithm for obtaining maximum likelihood estimates of the proportions of different strains in the sample. Using simulations, we show that the estimated proportions are close to the true proportions and that the phylogenetic imputation is highly accurate and improves the reference strains. We also apply this method to wastewater samples collected across the San Francisco Bay Area.

## Results

### Imputation

Many SARS-CoV-2 sequences submitted to public databases contain missing data (i.e., bases that are not coded as A, G, C, or T). This poses a problem when estimating the fraction of different SARS-CoV-2 strains, as strains with a high proportion of missing data in average will contain fewer nucleotide differences when compared to sequencing reads. We solve this problem using an imputation approach thereby allowing for a like-to-like comparison of reads against all reference strains. This method is in spirit similar to imputation approaches used in human genetics (e.g. ***Marchini and Howie, 2010***), although as we will show, due to the strong phylogenetic structure in the SARS-CoV-2 data, imputation is much more accurate than usually observed in diploid organisms. The method is based on calculating the posterior probability of each nucleotide in the leaf node of a phylogenetic tree and imputing based on the maximum posterior probability (see `Methods and Materials`). We compare the method (*Tree imputation*) to a naive imputation approach based on simply replacing missing nucleotides with the most frequent nucleotide observed in the alignment in that position (*Common allele imputation*). We evaluate the methods by first removing sequenced nucleotides in a real data set of 3,117,131 SARS-CoV-2 sequences and then re-imputing them using either *Tree imputation* or *Common allele imputation*.

For the vast majority of sites, *Tree imputation* has an error rate of *<* 5 × 10^−^4 although a few sites have imputation errors between 10^−^3 and 3 × 10^−^3 (Figure 1). The imputation error can be substantially higher for the naive *Common allele imputation* method with many sites showing error rates > 0.02 (Figure 1B). These are sites with high heterozygosity (Figure 1C) where substituting with the most common allele leads to high error rates. While the error rates for the *Common allele imputation* method naturally is predicted by the heterozygosity, the pattern is somewhat different for the *Tree imputation* method. The sites with highest imputation error are not the sites with highest heterozygosity, suggesting a high degree of homoplasy in these sites not directly predictable by the heterozygosity. These may be sites that switch allelic state often, i.e. have high mutation rates, but where the minor allele never increases substantially in frequency due to selection. An alternative explanation is sequencing errors. In fact, the site with the highest amount of apparent imputation error (position 24,410) is a site known to have a high proportion of sequencing errors (https://github.com/W-L/ProblematicSites_SARS-CoV2). It is located in a primer binding site where sequences containing the non-reference allele, A, often erroneously are assigned back to the reference allele, G, as a result of failed primer trimming during consensus building (https://github.com/W-L/ProblematicSites_SARS-CoV2). The A allele is one of the defining mutations of the delta strain and the apparent repeated re-emergence of the G allele within the delta clade (Figure S1) is likely a consequence of this common sequencing error. Most other sites, including the site with the highest heterozygosity, position 23,604 (Figure 1C), do not show a similar pattern of homoplasy (Figure S2). This suggests that the sites with the highest apparent imputation error rate, might in fact have a much lower true imputation error; the *Tree imputation* method may provide a more accurate assignment of alleles than the reported sequencing data for some problematic sequencing sites.

**Figure 1.**
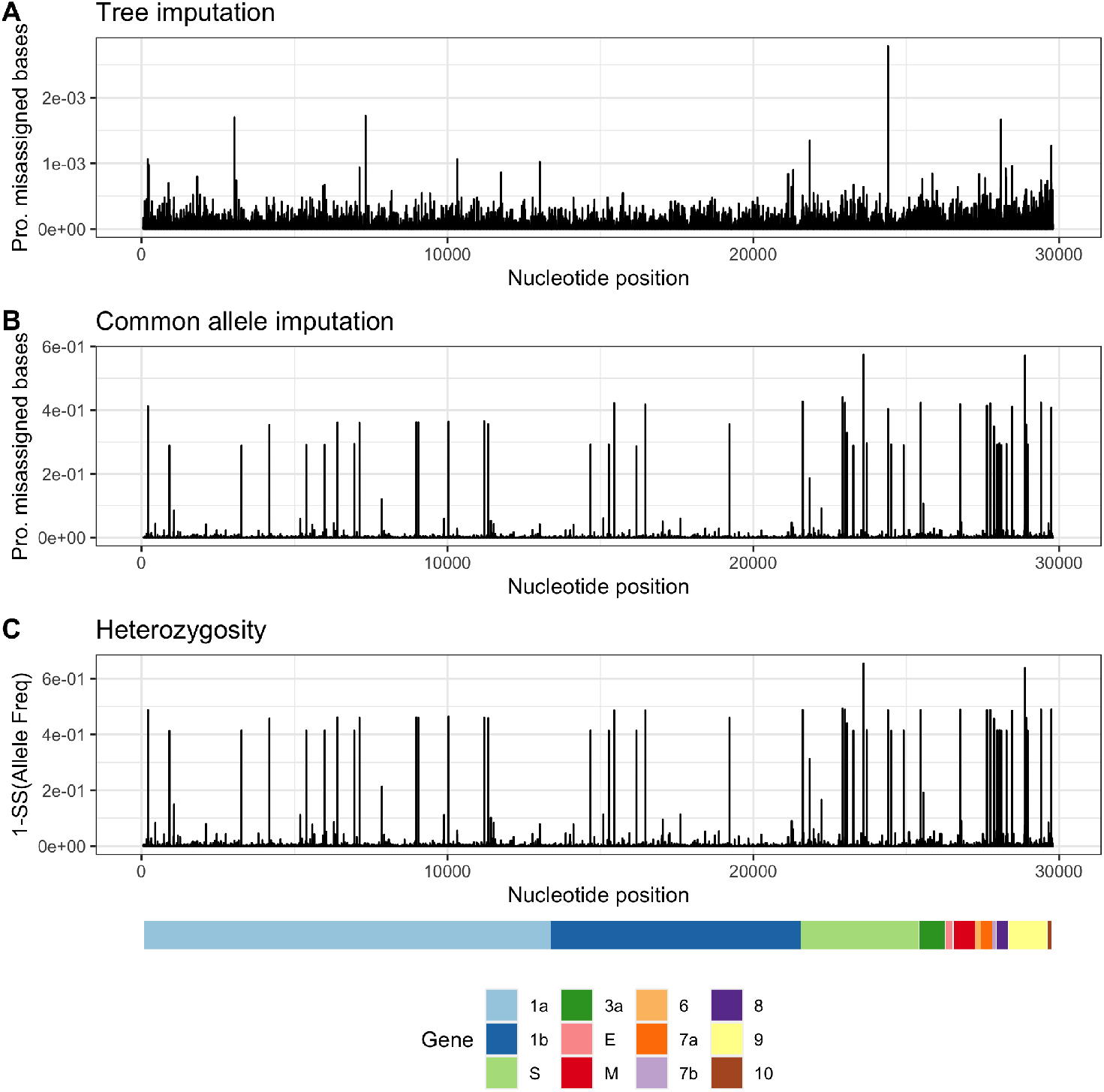
Proportion of missassigned bases along SARS-CoV-2 using the tree imputation method (A) and the common allele imputation method (B) against heterozygosity (C) using 3,117,131 SARS-CoV-2 genomes. Notice the difference in the scaling of the Y-axis of A and B.

### Simulations

In the Methods and Materials section, we describe an algorithm for estimating the proportion of different SARS-CoV-2 strains in an environmental sample using maximum likelihood. To evaluate the performance of the method, we simulate several sets of reads (single-end 300bp, paired-end 2×150bp, and paired-end 2×75bp) from 1, 3, 5, and 10 strains with an average depths of 100X, 500X, 1000X and a sequencing error rate of 0% and 0.5% (see `Methods and Materials`). We then apply the method to these sets of reads using a database of 3,117,131 strains and report the estimated proportions of each candidate strains and compare them with the truth (Figure 2, 3 and 4).

**Figure 2.**
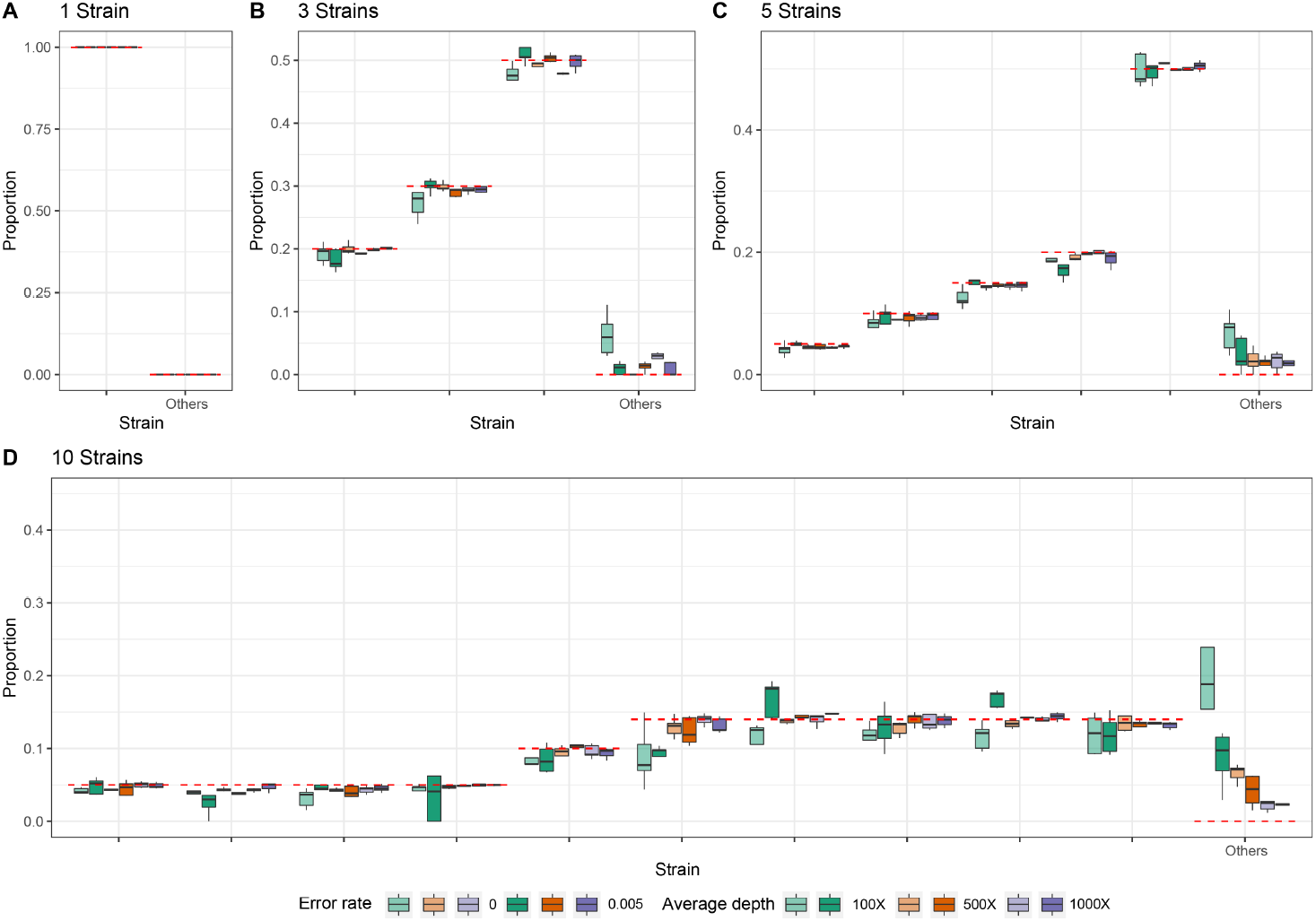
Estimated proportions for simulated 300 bp single-end reads with five replicates for when the sample truly contains 1 (A), 3 (B), 5 (C), or 10 (D) strains out of a total of 1,499,078 non-redundant candidate strains in the database. The red dashed lines indicate the true proportion of each strain. ‘Other’ indicates the sum of estimated proportions for all strains that are not truly represented in the sample.

**Figure 3.**
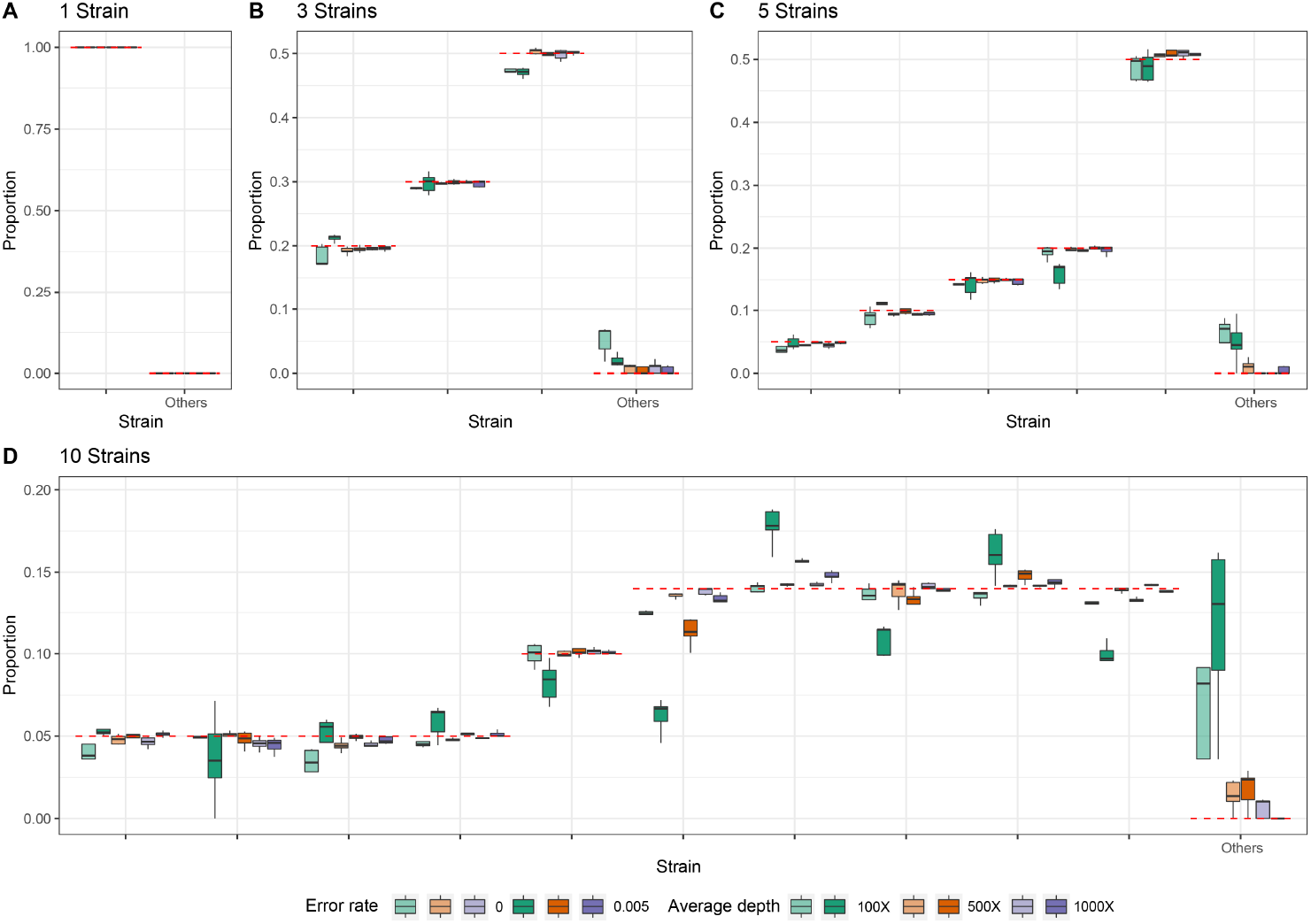
Estimated proportions for simulated paired-end reads (2×150 bp with an insert size of +25 bp) with five replicates for when the sample truly contains 1 (A), 3 (B), 5 (C), or 10 (D) strains out of a total of 1,499,078 non-redundant candidate strains in the database. The red dashed lines indicate the true proportion of each strain. ‘Other’ indicates the sum of estimated proportions for all strains that are not truly represented in the sample.

**Figure 4.**
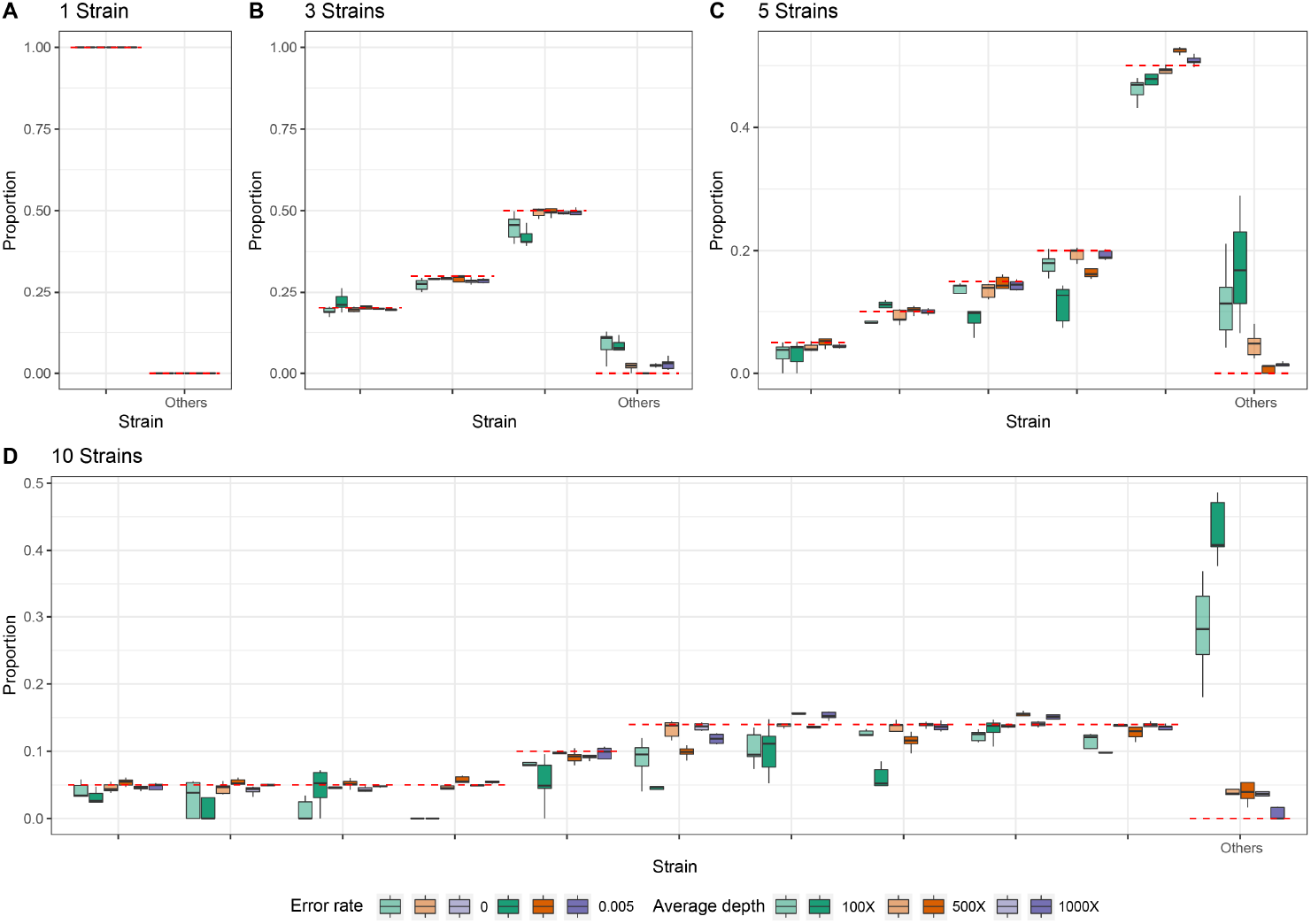
Estimated proportions for simulated paired-end reads (2×75 bp with an insert size of +25 bp) with five replicates for when the sample truly contains 1 (A), 3 (B), 5 (C), or 10 (D) strains out of a total of 1,499,078 non-redundant candidate strains in the database. The red dashed lines indicate the true proportion of each strain. ‘Other’ indicates the sum of estimated proportions for all strains that are not truly represented in the sample.

In most cases, the estimates are close to the true proportions, however, with a low coverage and high error rate, the proportions of the true strains will tend to be underestimated and strains that truly are not present will tend to be estimated as present in the sample. With one true strain in the sample, the proportion of this strain is always estimated to be 100%. For suffciently high depth, e.g. 1000*X* corresponding to roughly a total of 30 Mb of data, the estimates of strain proportions are quite accurate, even when 10 strains are present and for strains with a proportion as low as 5%. There is similarly very little probability mass assigned to strains that are not truly in the sample. For example, for 150 bp paired-end reads with a +25 bp insert and 1000X average sequencing depth, the estimate of the cumulative average proportion of all strains not truly in the sample is 0.63%.

The speed of the method is highly dependent on the number of true strains and the average depth (Figure 5), but for realistic sized data sets with a reference database of 3,117,131 strains, the typical computational time is between 15 minutes and two hours using a single core. This includes the initial time cost of ∼10.5 minutes for reading the large panel of reference strains into memory. There is no appreciable difference in speed between the different sequencing strategies used, except that paired-end 2×75bp sequences tends to take longer at higher average coverage. Simulations using the higher error rate (0.5%) are slower than simulations with no error. The average time for all sets of simulations with 5 or fewer true strains is <30 minutes for all coverages, while the average time for 10 true strains varies between ∼24 to ∼83 minutes depending on the average depth.

**Figure 5.**
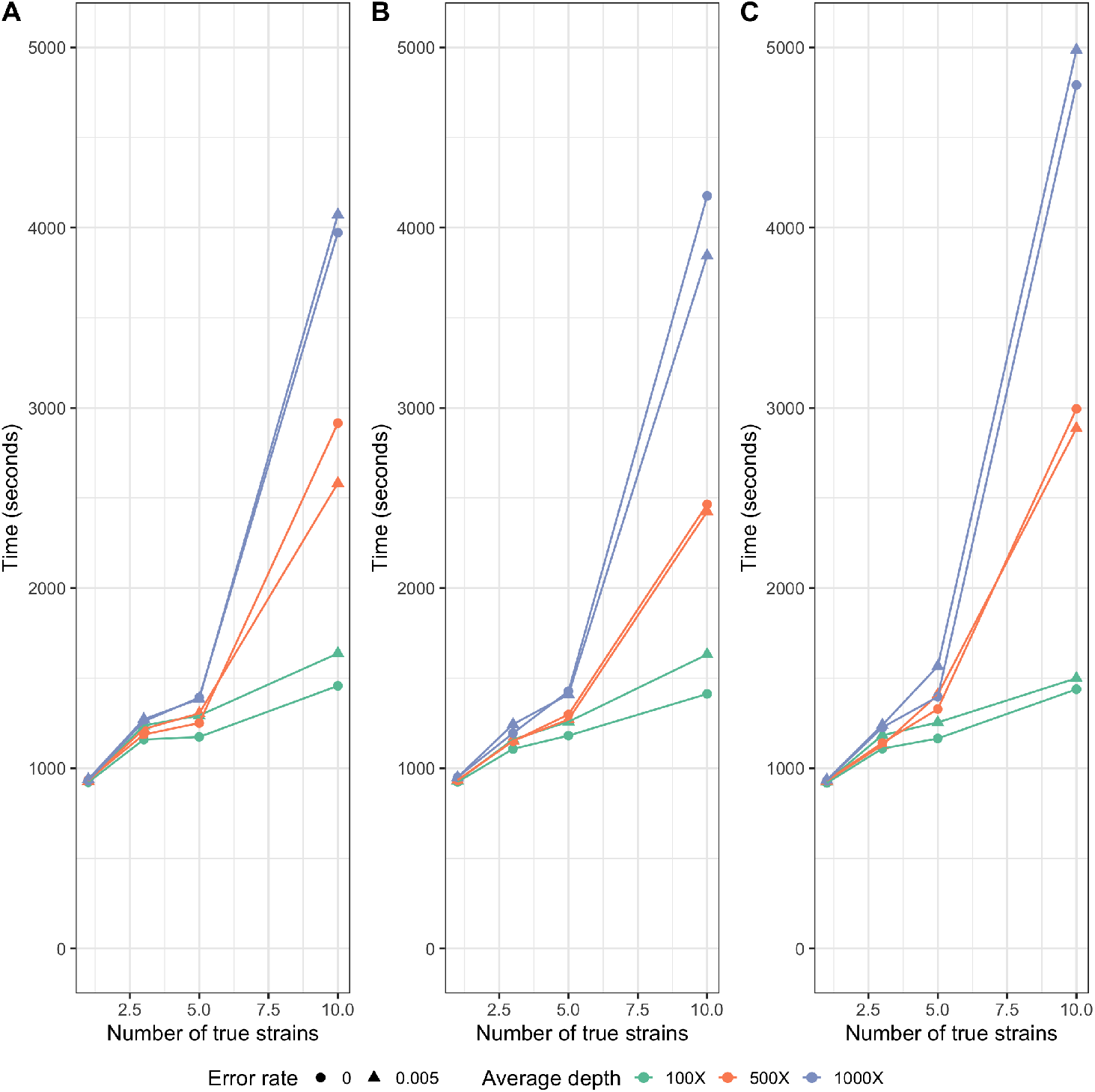
Average run times for single-end 300 bp (A), paired-end 2×150 bp (B), and paired-end 2×75 bp (C) read simulations using 100X, 500X, and 1000X average depth with an error rate of 0% and 0.5%. Each average run time reported is based on 5 replicates. Times were calculated using an AMD EPYC 7742 tetrahexaconta-core 2.25-3.40 GHz processor.

In order to quantify the statistical evidence for the presence of a candidate strain in the sample, we propose a likelihood ratio test, `LLR`, formed by comparing the maximum likelihood value calculated when the candidate strain is eliminated from the sample (*p* = 0) to the maximum likelihood value calculated when allowing the strain to be present in the sample (*p* ≥ 0), where *p* is the proportion of the strain in the sample (see `Methods and Materials`). Standard asymptotic theory for the distribution of the likelihood ratio statistics does not apply to this situation for several reasons, most importantly, a search is first made to find the strains that provide the largest increase in the likelihood among many strains, and we only calculate the likelihood ratio for the strains with estimates of *p* > 0. We, therefore, use simulations to evaluate the distribution of the likelihood ratio test statistics under varying conditions. We simulated 1,000 data sets with different numbers of true strains, coverage, read length and error rate and calculated the likelihood ratio for all strains that were falsely inferred to be present in the sample (Figure 6). Since the frequency of `LLR` > 2 and `LLR` > 4 is about 0.001 and 0.0005, respectively, we recommend using 2 and 4 as thresholds for strong and extremely strong evidence for presence of the strain in the sample.

**Figure 6.**
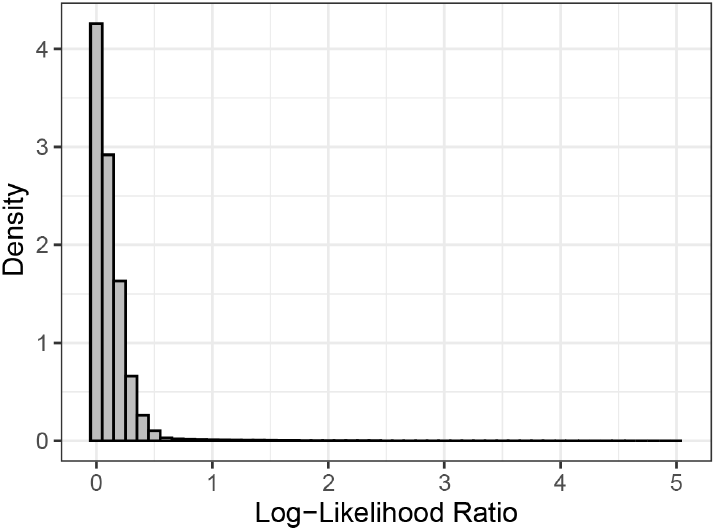
Distribution of log-likelihood ratios from 1,000 data sets of simulated 100 ∼ 300*bp* single-end reads. Those simulated data sets include 3 ∼ 10 strains with proportions ranging from 5% to 50% and average depths ranging from 50X to 300X. The error rate varies from 0% to 0.5%.

### Application to wastewater data from *Crits-Christoph et al. (2021*)

To apply our method to a published data set, we estimated the composition of SARS-CoV-2 lineages using wastewater shotgun sequencing data from ***Crits-Christoph et al. (2021***) in Figure 7, which were all collected in the San Francisco Bay Area. Two out of the top ten strains were collected in Alameda county (EPI_ISL_625508, which is identical to EPI_ISL_625520, and EPI_ISL_672326), and the top five strains were all collected in North America.

**Figure 7.**
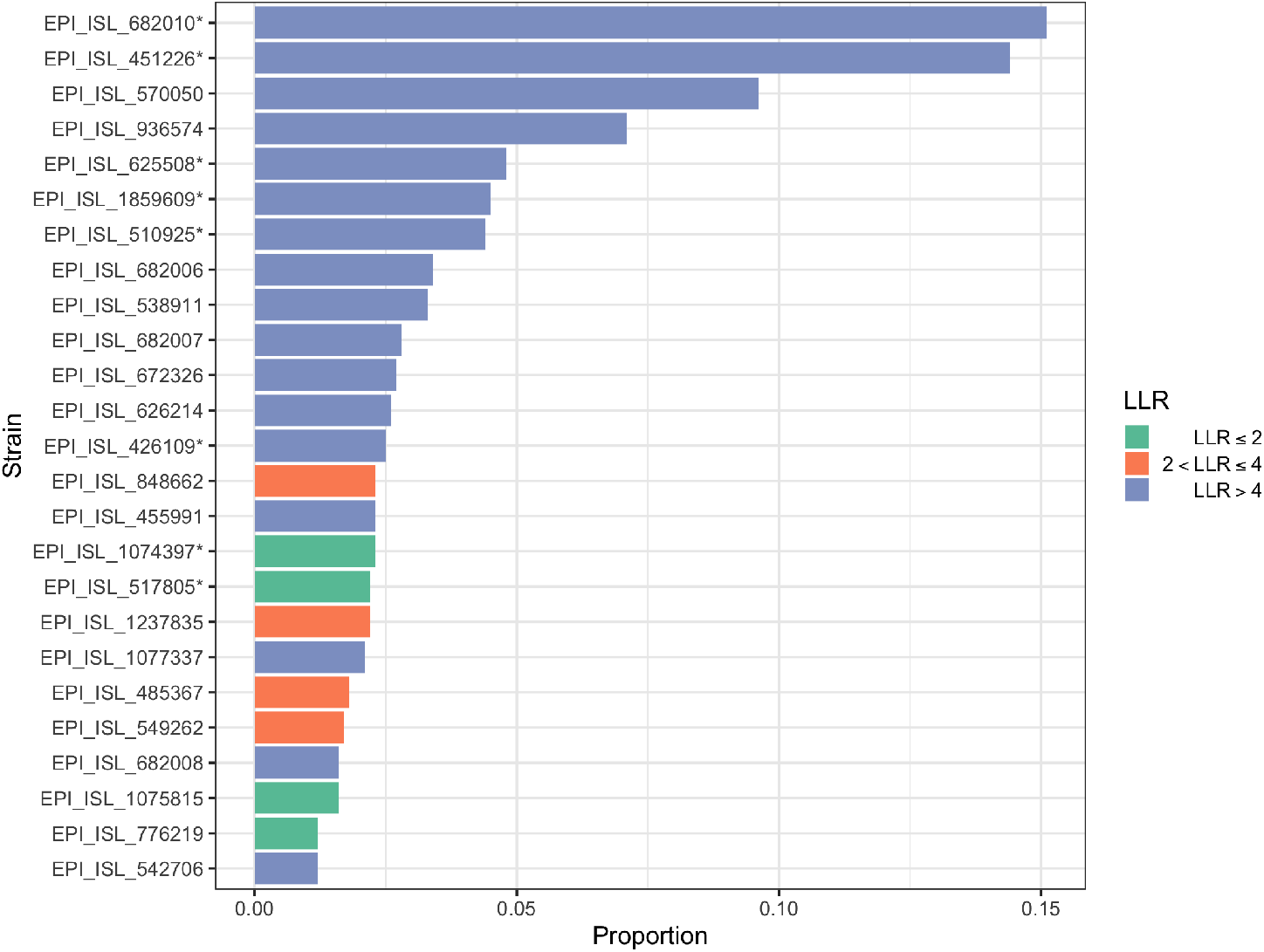
Estimated proportions of the top 25 strains estimated from wastewater shotgun sequencing data from ***Crits-Christoph et al. (2021***) and their log-likelihood ratios. Strains with an asterisk (*) are identical with other strains. EPI_ISL_682010* is identical to EPI_ISL_682025, EPI_ISL_1373628, EPI_ISL_1373632, and EPI_ISL_1373659. EPI_ISL_451226* is identical to EPI_ISL_451227 and EPI_ISL_455983. EPI_ISL_625508* is identical to EPI_ISL_625520, EPI_ISL_672318, EPI_ISL_672449, EPI_ISL_739003, EPI_ISL_739029, EPI_ISL_739135, EPI_ISL_739161, EPI_ISL_739207, and EPI_ISL_739286. EPI_ISL_1859609* is identical to EPI_ISL_1859762. EPI_ISL_510925* is identical to EPI_ISL_510926. EPI_ISL_426109* is identical to EPI_ISL_486012, EPI_ISL_570168, EPI_ISL_570172, EPI_ISL_576500, and EPI_ISL_576501. EPI_ISL_1074397* is identical to EPI_ISL_2190584. EPI_ISL_517805* is identical to EPI_ISL_527398 and EPI_ISL_137362.

## Discussion

In order to allow for accurate inferences of strain composition, we first developed a new phylogenetic method for data imputation for SARS-CoV-2 sequences. The method proved to be highly accurate with error rates comparable to, or lower, than typical sequencing error rates (Figure1A). In fact, apparent wrongly inferred nucleotides may in many cases not be wrongly inferred but rather be inferences of the true allele, correcting a sequencing error in the reported sequence. Thus, similarly to imputation-based genotype calling in humans, this method could be used for correcting sequencing errors and incorporated formally into an algorithm of imputation-informed sequencing where the quality scores from sequencing reads are combined with phylogenetically informed nucleotide probabilities to call nucleotides in each position. Computationally, this could be done simply by using the phylogenetic posterior probabilities of nucleotides as priors for genotype calling.

Our simulation results for the EM algorithm show that the new method can accurately estimate proportions of SARS-CoV-2 lineages in wastewater samples when up to 10 strains with frequencies as low as 5% are represented in the sample. Nonetheless, the estimated proportions for the true strains tend to be slightly lower than the actual proportions because the presence of other non-true strains is also estimated at alow frequency. In order to have some probability for other non-true strains to be estimated, the true proportions for the true strains will naturally in average be slightly underestimated. In all sets of simulations of single-end 300bp reads (Figure 2), paired-end 2 × 75bp (Figure 3), and paired-end 2 × 150bp (Figure 4), the estimated proportions of the true strains tend to be more accurate as sequencing depth increases. When there are many strains (i.e., when there are 10 true strains) and sequencing depth is low (i.e., 100X), there is a high degree of noise in the data set. However, as the total sequencing depth increases, the estimates become progressively more accurate. We recommend that studies focused on identifying different strains of SARS-CoV-2 in environmental samples aim to achieve an average depth of 1000X. Additionally,the method presented here has only been evaluated for the estimation of proportions of strains with a frequency of 5% or larger. We recommend that strains identified in the sample at low frequencies are evaluated using the likelihood ratio test as they likely could be false positives.

Current strategies for monitoring community composition of SARS-CoV-2 strains include sequencing a large number of clinical samples. As SARS-CoV-2 becomes endemic, tracking the relative prevalence in local communities of different SARS-CoV-2 strains will be highly costly. Furthermore, the use of clinical samples is associated with a lag from infection onset to hospitalization. Our results suggest an alternative strategy of monitoring using wastewater samples. Wastewater sequencing has already proved effective for tracking SARS-CoV-2 abundance ((***Korber et al., 2020***; ***Rockett et al., 2020***)). With the computational framework developed here, it also promises to become an important cost-effective strategy for monitoring the local composition of different viral strains.

## Methods and Materials

### SARS-CoV-2 Reference Database

To build the SARS-CoV-2 reference database, a multiple sequence alignment (MSA) of 3,117,131 SARS-CoV-2 genomes (msa_2021-10-15.tar.xz) and the corresponding phylogenetic tree (GISAID-hCoV-19-phylogeny-2021-10-13.zip) was downloaded from GISAID (www.gisaid.org) on October 16, 2021. We pruned sequence EPI_ISL_4989640 from the tree since it was not present in the MSA. We use the function `collapse.singles` to collapse elbow nodes (i.e., nodes other than the root with two degrees) and `multi2di` to resolve multichotomies in the **R** `ape` package (***Paradis et al., 2004***). We impute missing data (i.e., every position in the MSA that did not contain an A, G, C, or T), using the phylogenetic tree. To do so, we first scale the branch lengths in terms of substitutions per site by dividing each reported branch length by the average sequence length (29618.5). For branch lengths that were reported to be 0, we define them to be 0.01 divided by the average sequence length. We impute missing nucleotides using the maximum of the posterior probability of each nucleotide in the leaf nodes under a standard Jukes and Cantor model (***Jukes et al., 1969***), using standard computational algorithms (***Yang, 2014***). In brief, because the model is time-reversible, the root can be placed in any particular node, and the fractional likelihoods (joint probabilities of a fraction of the data in the leaf nodes and the nucleotide state in the node) can be pulled recursively towards the node from both the child nodes and the parental node. The posterior probability in the leaf nodes of a nucleotide is calculated as the product of the stationary probability of the nucleotide multiplied by the fractional likelihood in the leaf node conditioned on the data in all other leaf nodes. This can be programmed so the calculation is linear in the number of leaf nodes using a single pre-order and a single post-order traversal of the tree that will calculate the posterior probabilities in all nodes. We note that other models than the Jukes and Cantor model could provide more accurate estimates, but at a computational cost.

Since calculating fractional likelihoods for the entire tree requires more RAM than was computationally feasible for us (∼72TB of RAM), we split the tree into partitions, and process each partition sequentially as follows:

Each internal node in the tree corresponds to a partition of leaf nodes into three sets. First, we identify the node with the minimum variance in the number of elements among these three partitions, i.e. we find

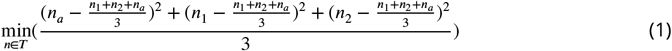

where *n* is a node in the tree, *T* is the tree, *n*_1_ is the number of leaf nodes descending from the left child of *n, n*_2_ is the number of leaf nodes descending from the right child, and *n*_*a*_ = *N* − *n*_1_ − *n*_2_, where *N* is the total number of leaf nodes in the tree. We then split the tree into 3 subtrees by eliminating the identified node. We then iterate this procedure for the resulting subtrees until all trees contain at most 50,000 leaf nodes.

Using this partitioning procedure, we obtain 121 trees which we use to calculate the posterior probabilities at each site. After imputation, we trim the MSA to begin at the start of the Wuhan reference sequence (Wuhan-Hu-1), position 55 in the MSA, and we removed every position in the MSA that contains a gap in Wuhan-Hu-1. After this trimming and imputation process, we save non-informative invariant sites (856 sites), in order to reduce running time when eliminating unlikely strains. We also remove all identical sequences, resulting in 1,499,078 non-redundant genomes.

### Estimating the proportions of SARS-CoV-2 genomes

All sequencing reads are aligned to Wuhan-Hu-1 (NC_045512.2) using bowtie2 (***Langmead and Salzberg, 2012***) with the following command for single-end reads, `bowtie2 –all -f -x wuhCor1 -U`, and for paired-end reads, `bowtie2 –all -f -x wuhCor1 −1 −2`. For each read data set, we first remove unlikely genomes from the candidate strain alignment by eliminating genomes with SNP alleles that have an allele frequency in the read data less than a user-defined frequency threshold. For the analyses in this data, that threshold was set to 0.01. This typically reduced the size of the alignment to *<* 1, 000 relevant genomes. Using this reduced set of SARS-CoV-2 genomes, we calculate a matrix of dimensions (number of reads)×(number of genomes) containing the number of mismatches between each sequencing read and each genome, *d* = {*d*_*ij*_}. For paired-end reads with reads that overlap, we use the consensus nuleotide. If there is a conflict at any position in the overlap of the paired-end reads, we omit this site. Based on the mismatch matrix, *d*, we first calculate the probability of observing read *j* given that it comes from strain *i*, denoted as *q*_*ij*_. Assuming that the reads are independent (PCR clones removed) and a user-defined error rate *α* (default = 0.005) at each nucleotide, this probability is given by

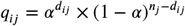

where *n*_*j*_ is the length of read *j* and *d*_*ij*_ is the number of mismatches in read *j* given that it comes from strain *i*. The log-likelihood is then given by

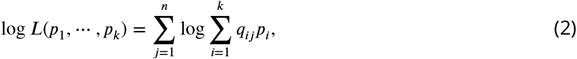

where *p*_*i*_ (*i* = 1,., *k*) is the proportion of strain *i*, i.e. the parameters we wish to estimate. We then use the standard Expectation Maximization (EM) algorithm (***Dempster et al., 1977***) to maximize the likelihood function with respect to these parameters 1:

#### Algorithm 1 EM algorithm for estimating the proportions of candidate strains

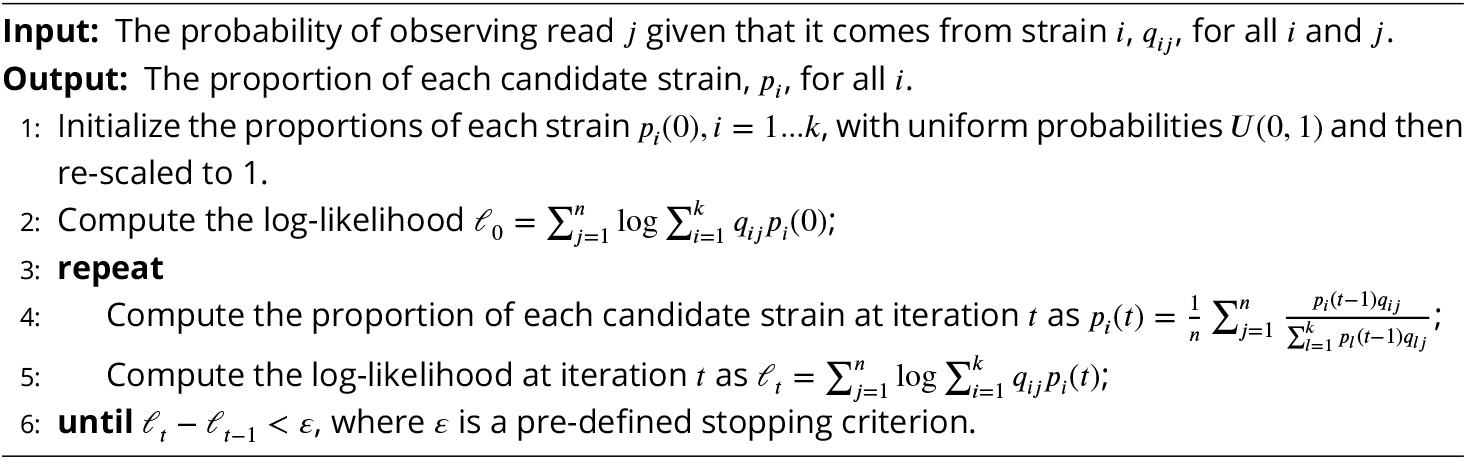

However, Algorithm 1 usually has a slow convergence rate, especially when the number of candidate strains *k* is large. Therefore, to accelerate the Algorithm 1, we use the SQUAREM algorithm proposed by ***Varadhan and Roland (2008***) with its implementation in the **R** package `turboEM` (***Bobb and Varadhan, 2020***).

### Determining unidentifiable strains

Note that if two stains have the same *q*_*ij*_’s, say there exist *i* and *i*′ such that *q*_*ij*_ = *q*_*i′j*_ for all *j* = 1,…., *n*, the log-likelihood (2) becomes

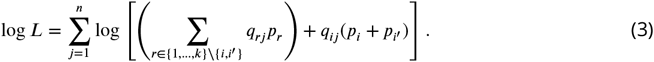

Therefore, as long as *p*_*i*_ + *p*_*i′*_ is fixed, (3) remains the same no matter what value *p*_*i*_ and *p*_*i′*_ take, making the model unidentifiable. To solve this problem, we gather strains with the same 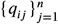 into an unidentifiable group and estimate its overall proportion instead of the proportions of each strain in it.

### Quantifying the statistical evidence of the existence of each candidate strain

To provide a measure of statistical support for the presence of strain *i*_0_, i.e.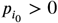, we remove strain *i*_0_ from the candidate set of strains and re-run Algorithm 1 providing a new estimate 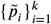 with 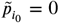. Using (2), we can then calculate the difference in log likelihood before and after removing strain *i*_0_, denoted as 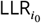. From our simulations (see Results), we recommend using 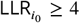 as strong statistical evidence in favor of existence of strain *i*_0_ in the sample.

### Simulating missing data for imputation

For every SARS-CoV-2 genome (out of a total of 3,117,131 genomes), we randomly remove 1% of nucleotides, and save the true nucleotide at each position that was removed. We then use the *Tree imputation* method and the *Common allele* method to impute the nucleotides that are missing.

### Simulating reads from SARS-CoV-2 genomes

We choose 10 strains among 1,499,078 strains uniformly at random. Then, to simulate single-end reads from a strain, we choose a starting point uniformly at random and let it extend *m*_0_ bps, where *m*_0_ is the read length. For paired-end reads, we similarly choose a starting point at random and let it extend *m*_0_ bps. Then, starting from the end of this read, if the insert size is *m*_1_ is positive, we simulate the start of the reverse read *m*_1_ bps forward with length *m*_0_; if *m*_1_ is negative, we simulate the start of the reverse read *m*_1_ bps backwards. We then add sequencing errors independently with probability *α* = 0.005 at each site. Errors are induced by relabeling the nucleotide to any of the other three possible nucleotides with the following probabilities used in ***Stephens et al. (2016)***:

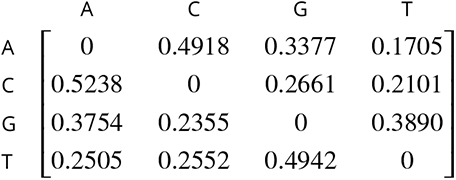

### Calculating time cost

To calculate running time of the method we use `/usr/bin/time` on an AMD EPYC 7742 tetrahexaconta-core 2.25-3.40 GHz processor and report `real time` in the results (Figure 5). The running time that we calculate includes running the method from start (reading in the reference strains) to finish (reporting proportions) and includes the filtering step for eliminating unlikely strains. We report times that do not include calculating the log-likelihood ratio.

### Applying the method to wastewater data from *Crits-Christoph et al. (2021*)

Wastewater shotgun sequencing data from ***Crits-Christoph et al. (2021***) was downloaded from NCBI BioProject ID PRJNA661613 (https://www.ncbi.nlm.nih.gov/bioproject/?term=PRJNA661613). All samples were pooled together and aligned against Wuhan-Hu-1 using BWA-MEM (***Li, 2013***) to identify SARS-CoV-2 reads.

## Data Availability

Simulations used in this manuscript can be downloaded at https://doi.org/10.5281/zenodo.5838942. The imputed MSA can be downloaded at https://doi.org/10.5281/zenodo.5838946. Identical strains are contained in the headers of the MSA separated by colons. Software for the method is available for download at https://github.com/lpipes/SARS_CoV_2_wastewater_surveillance.

https://doi.org/10.5281/zenodo.5838946

https://github.com/lpipes/SARS_CoV_2_wastewater_surveillance

## Acknowledgments

We gratefully acknowledge all laboratories who submitted SARS-CoV-2 genome sequences to the GISAID EpiCoV database (www.gisaid.org), which we used for the reference database for this method. We acknowledge Xiaoyi Gu for testing the software and for development of a website portal for the method, and Selina Kim for working on this project.

## Supplementary Material

**Figure S1.**
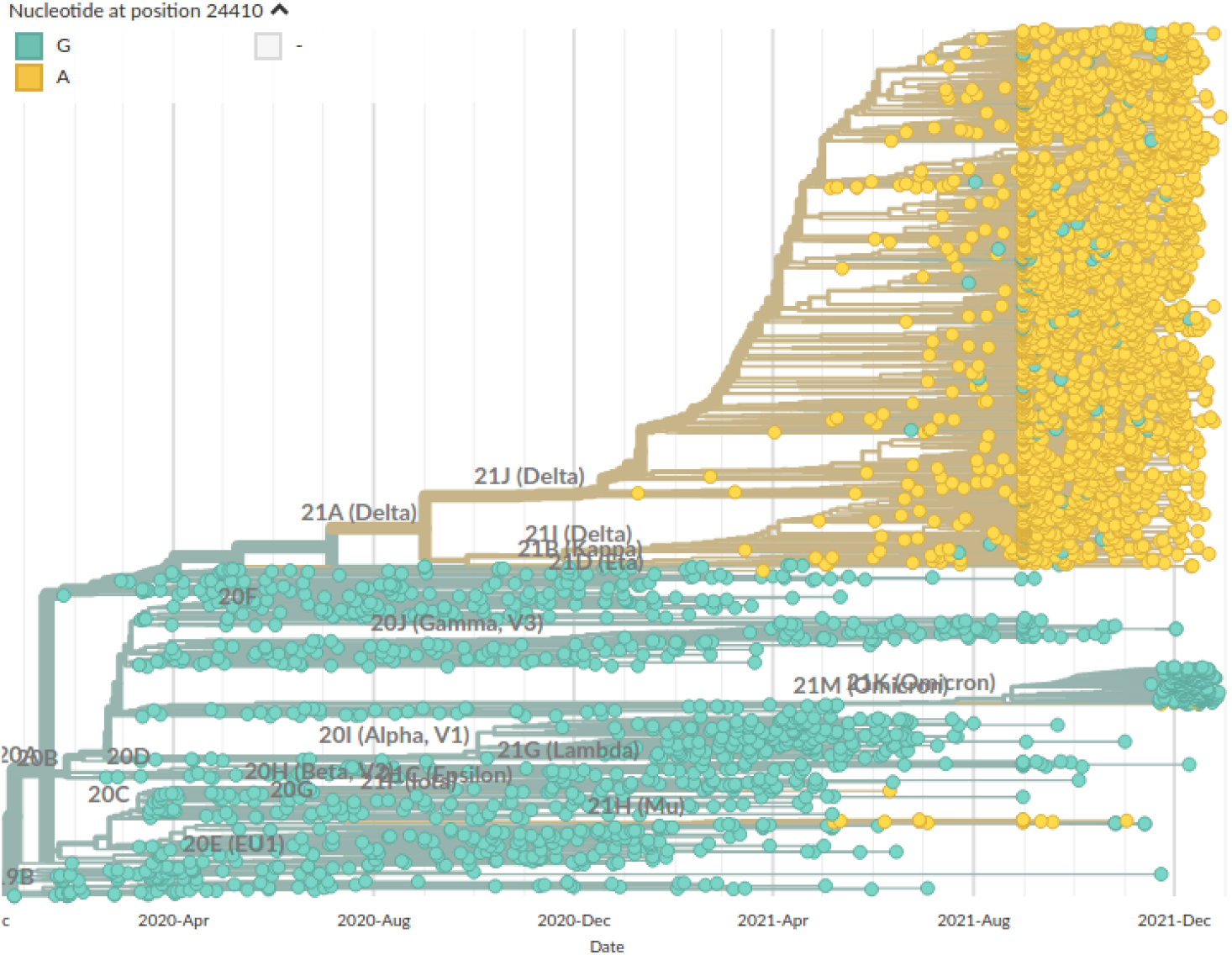
Screenshot of SARS-CoV-2 phylogeny from nextstrain.org for nucleotide position 24,410 taken on January 5, 2022.

**Figure S2.**
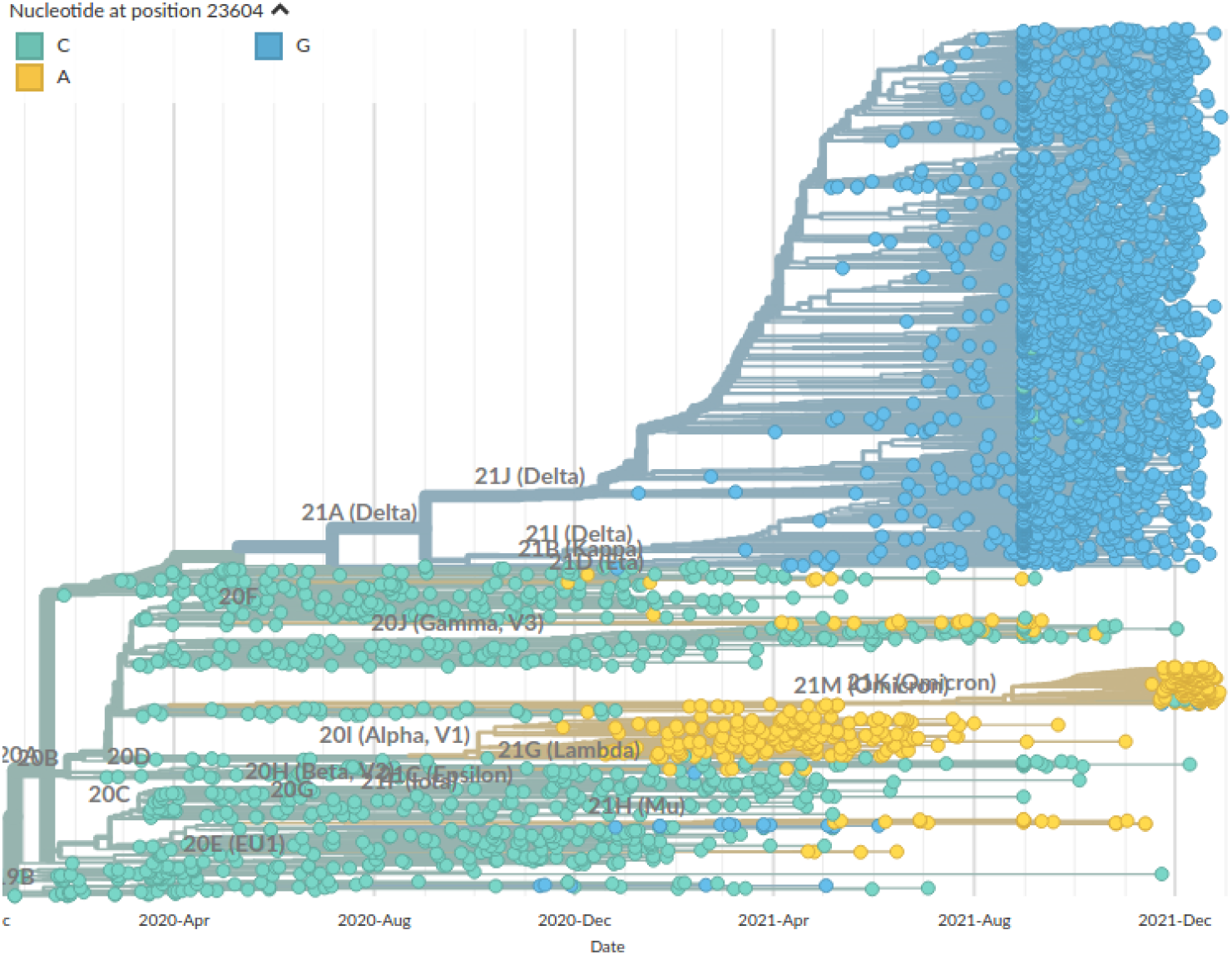
Screenshot of SARS-CoV-2 phylogeny from nextstrain.org for nucleotide position 23,604 taken on January 5, 2022.

